# A beta-Poisson model for infectious disease transmission

**DOI:** 10.1101/2023.03.24.23287701

**Authors:** Joe Hilton, Ian Hall

**Affiliations:** School of Life Sciences and Zeeman Institute (SBIDER), University of Warwick, Coventry, UK; Department of Mathematics and School of Health Sciences, University of Manchester, Manchester, UK

## Abstract

Outbreaks of emerging and zoonotic infections represent a substantial threat to human health and well-being. These outbreaks tend to be characterised by highly stochastic transmission dynamics with intense variation in transmission potential between cases. The negative binomial distribution is commonly used as a model for transmission in the early stages of an epidemic as it has a natural interpretation as the convolution of a Poisson contact process and a gamma-distributed infectivity. In this study we expand upon the negative binomial model by introducing a beta-Poisson mixture model in which infectious individuals make contacts at the points of a Poisson process and then transmit infection along these contacts with a beta-distributed probability. We show that the negative binomial distribution is a limit case of this model, as is the zero-inflated Poisson distribution obtained by combining a Poisson-distributed contact process with an additional failure probability. We assess the beta-Poisson models applicability by fitting it to secondary case distributions (the distribution of the number of subsequent cases generated by a single case) estimated from outbreaks covering a range of pathogens and geographical settings. We find that while the beta-Poisson mixture can achieve a closer to fit to data than the negative binomial distribution, it is consistently outperformed by the negative binomial in terms of Akaike Information Criterion, making it a suboptimal choice on parsimonious grounds. The beta-Poisson performs similarly to the negative binomial model in its ability to capture features of the secondary case distribution such as overdispersion, prevalence of superspreaders, and the probability of a case generating zero subsequent cases. Despite this possible shortcoming, the beta-Poisson distribution may still be of interest in the context of intervention modelling since its structure allows for the simulation of measures which change contact structures while leaving individual-level infectivity unchanged, and vice-versa.

## Introduction

Infections at the human-animal interface are often associated with high levels of morbidity and mortality, making them a subject of substantial interest to mathematical modellers and other infectious disease researchers [1]. These infections often have low reproductive ratios, meaning that their behaviour is characterised by “stuttering” outbreaks, with each case infecting only a small number of contacts and local extinctions occurring frequently [1–3]. These dynamics are also characteristic of outbreaks of infections such as measles and mumps in populations which are mostly vaccinated and where the infection is close to eradication [1, 2]. Stuttering outbreaks with low numbers of cases permit more complete observation than larger epidemics and pandemics, allowing public health workers and researchers to identify individual transmission events and estimate a *transmission chain*, the network obtained by drawing an edge from each case back to its infector. The topology of this network characterises the epidemic, with its mean giving the basic (in the case of a novel infection with no preexisting immunity) or effective (in the close-to-eradication setting) reproductive ratio [2].

The key to interpreting transmission chain behaviour from a mathematical perspective is the theory of *branching processes*. Branching process models provide a rigorous description of an outbreak’s early behaviour in the stages before the underlying susceptible population becomes depleted [4] and are a well-established item in the inventory of mathematical methods available to epidemiologists [1, 3, 5–10]. A branching process model can be formulated from transmission chain data by using the degree distribution of the transmission network as the branching process’s *offspring distribution*, the distribution of the number of cases generated by each case. We refer to these cases as *secondary cases*. In some cases rather than a fully described transmission chain we may just know the number of secondary cases assigned to each case in the outbreak, and we will refer to such data as *secondary case data*. Results from the theory of branching processes can be used to calculate useful quantities like the outbreak size distribution and probability that an outbreak goes extinct [11]. One of the major benefits of branching process models over more typical compartmental models is that they are not tied to the combination of a Poisson contact process and exponential infectious period which underlies most compartmental modelling. Although models for infections with non-exponential infectious periods can be simulated and analysed, they are in general non-Markovian and call for sophisticated mathematical techniques [12]. In particular, branching processes whose offspring distribution is the same at every generation, known as Galton-Watson processes, are always Markovian since the number of cases in a given generation depends only on the number in the last generation and are easily simulated on this generation-by-generation basis by drawing successive random numbers from the offspring distribution.

While some studies use the empirical transmission chain degree distribution as an offspring distribution [8, 13], it is more common to use a parametric model. Commonly used offspring distributions include the Poisson [5], geometric [14], and negative binomial [2, 3, 9, 15] distributions. The geometric and negative binomial models are both examples of *mixed Poisson distributions* [16]. These are Poisson distributions where the Poisson parameter is allowed to vary according to some continuous mixing distribution - in the case of the geometric and negative binomial, an exponential and a gamma distribution respectively. The branching process model with geometric offspring distribution specifically captures the early behaviour of the homogeneous stochastic SIR model, where the exponentially-distributed infectious period and Poisson process contact behaviour combine to give an exponential-Poisson mixture. Mixed Poisson distributions are always *overdispersed*, meaning their variance is larger than their mean, in contrast to the (unmixed) Poisson, whose mean and variance are equal [16].

The negative binomial model’s overdispersion gives it the capacity to model superspreading events, where the number of secondary cases generated by a single case is substantially more than the mean. Lloyd-Smith *et al*. developed a more formal definition, where a superspreading event for an infection with effective reproductive ratio *R* is a transmission event in the upper *n*th percentile (in their study they take *n* = 99) of the Poisson distribution with mean *R* [2]. Examples of superspreading have been recorded in novel coronavirus outbreaks [17, 18], and in the close-to-eradication setting in measles [19].

Another approach to modelling overdispersed count data is to use *zero inflation*. Zero inflated distributions specifically model count data with more zeros than would be expected under ordinary modelling assumptions [20, 21]. This is a natural interpretation to consider since transmission chains often contain a high proportion of individuals who do not produce any subsequent cases. In this context zero inflation can be interpreted as modelling situations in which infectious cases are unable to engage in ordinary contact behaviour either because of hospitilisation and effective control measures, or simply because they are too unwell.

In this article we introduce an approach which models the number of secondary cases produced by an infectious individual as a beta-Poisson mixture, a Poisson distribution whose mean is scaled by a beta-distributed random variable. Early formulations of the beta-Poisson distribution date back to the 1960’s [22], and in infectious disease modelling contexts it is sometimes used as a dose-response model [23, 24]. However, it is not typically used as a person-to-person transmission model, despite a natural and intuitive interpretation as the combination of a social contact formation process and a transmission process across those contacts. Under this interpretation, each case is assigned a transmission probability drawn from a beta distribution, and makes a Poisson-distributed number of contacts during their infectious period. The beta-distributed transmission probability tells us the probability that a given social contact results in a successful transmission event. The distribution of transmission probability is intended to capture individual-level differences in contact behaviour (for instance, the high frequency of physical touch-based contacts made by children relative to adults [25]) and physiological response (such as age-dependent effects in COVID-19 symptomaticity [26]). We will assess the effectiveness of the beta-Poisson mixture as a transmission model by fitting it to eight sets of secondary case data and compare its performance with the Poisson, geometric, negative binomial, and zero-inflated Poisson (ZIP) distributions.

## Results

Code for replicating the following analysis is available at

github.com/JBHilton/beta-poisson-epidemics.

The beta-Poisson model has three parameters: the mean number of secondary cases *λ*, a measure of overdispersion Φ, and the mean number of contacts *N* made over an individual’s infectious period. Mechanistically, it describes the following transmission process: each infectious individual draws a transmission probability *p* from a beta distribution with parameters *α*_1_ = *λ*Φ and *α*_2_ = (*N* − *λ*)Φ, draws a number of contacts from a Poisson distribution with mean *N*, and infects each of these contacts with probability *p*. The total number of contacts generated by an individual with infection probability *p* is therefore Poisson with mean *pN*. This is a generalisation of more standard definitions of the beta-Poisson distribution [22–24], where the Poisson parameter itself is *p*, equivalent to the specific case *N* = 1 in our model.

In the Methods section we demonstrate that four other common choices of secondary case distribution (Poisson, geometric, negative binomial, and zero-inflated Poisson) [2] can be considered special limiting cases of the beta-Poisson. In the limit *N* ⟶ ∞, the beta-Poisson distribution is equivalent to the negative binomial distribution with mean *λ* and overdispersion *θ* = Φ^−1^. Because the negative binomial distribution has the Poisson and geometric as special cases (overdispersion *θ* = 0 and *θ* = *λ* respectively) these special cases are inherited by the beta-Poisson. The zero-inflated Poisson distribution is a convex combination of a Poisson distribution with mean 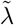 and a point probability mass at zero with convexity parameter *σ*, so that the probability of obtaining zero is given by *σ* plus the probability of drawing zero under the Poisson distribution, and the probability of obtaining *x >* 0 is given by the Poisson probability of obtaining *x* multiplied by (1 − *σ*). The natural epidemiological interpretation of this distribution is that individuals either generate a Poisson number of secondary cases with probability 1 − *σ*, or are prevented from making contacts due to either illness or a planned intervention and do not generate secondary cases with probability *σ*. This is analogous to our beta-Poisson distribution in the limit Φ ⟶ 0 (i.e. *α*_1_ + *α*_2_ ⟶ 0). In this limit the underlying beta distribution tends to a two-point distribution with probability *α*_2_*/*(*α*_1_ + *α*_2_) at zero and probability *α*_1_*/*(*α*_1_ + *α*_2_) at one, so that the beta draw tends towards the choice in the ZIP distribution between generating zero cases or generating a Poisson-distributed number of case. The ZIP limit of the beta-Poisson distribution will have Poisson parameter 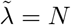 and zero inflation parameter *σ* = 1 − *λ/N*. A visual comparison of the mechanisms underlying the different offspring distribution models is provided in Fig 1. To establish whether or not the beta-Poisson offers a meaningful improvement over these limiting distributions, we will fit all five models to transmission chain data and compare their fitting behaviour.

**Fig 1.**
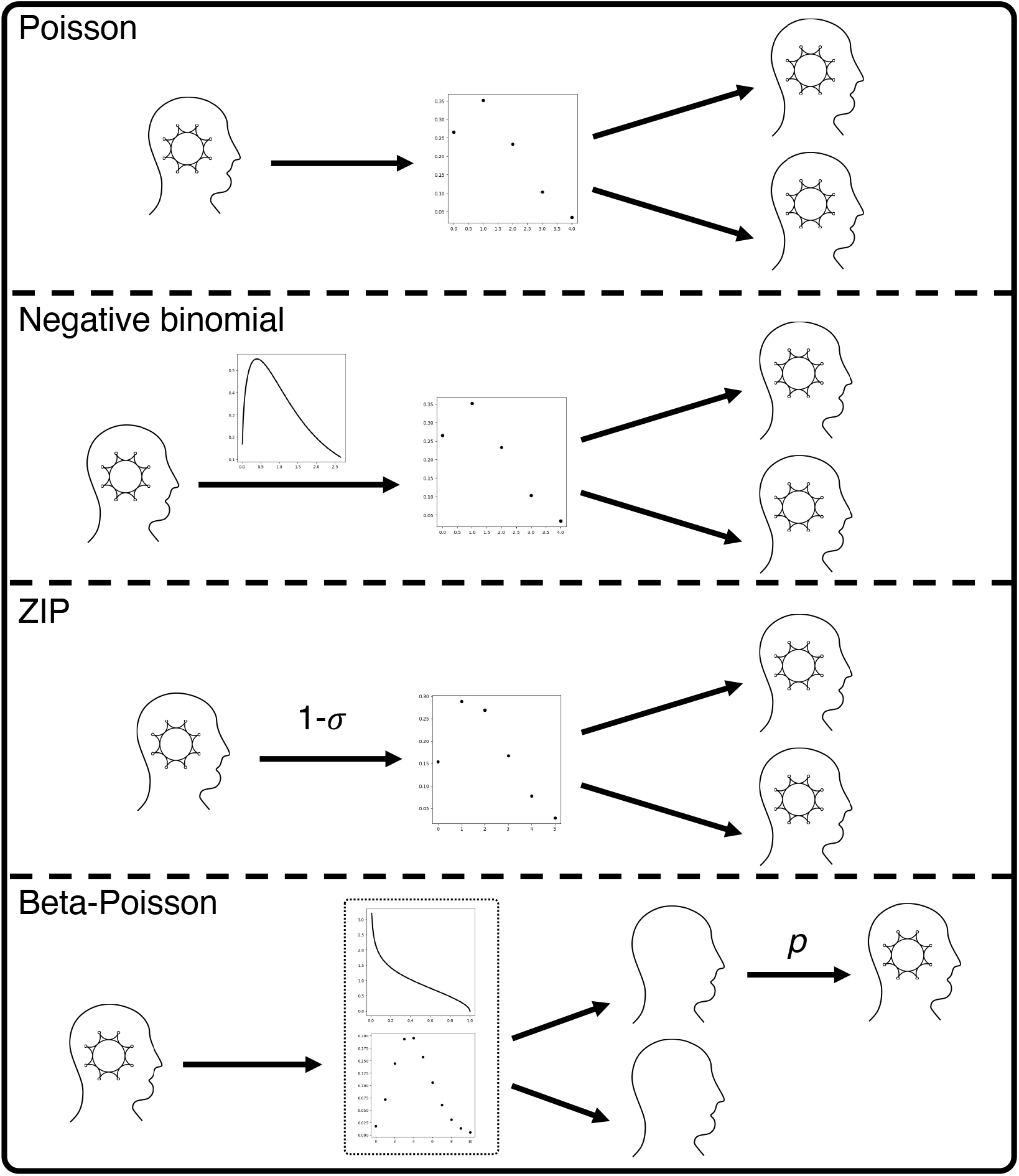
Mechanistic comparison of the different offspring distribution models. Under a Poisson model, each infectious individual generates a number of secondary cases, drawn from a Poisson distribution which is the same for all infectious individuals. Under a negative binomial model, each infectious individual is assigned an innate infectivity, drawn from a gamma distribution, which defines a Poisson distribution from which they draw their secondary cases. Under the zero-inflated Poisson model, each infectious individual either generates no secondary cases with probability *σ*, or else with probability 1 − *σ* generates a Poisson-distributed number of secondary cases, with the same Poisson parameter for all infectious individuals. Finally, under the beta-Poisson distribution each infectious individual is assigned their own infection probability *p* from a beta distribution, makes a Poisson-distributed number of contacts, and then infects each of these contacts with probability *p*.

For the Poisson and geometric models 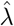, the MLE of *λ*, is given by the sample mean. We calculate confidence intervals for these model fits using a grid calculation. For the negative binomial model we find 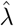 by calculating the sample mean and calculate the MLE of *θ* by finding the maximum of its log likelihood function numerically using the scipy.optimize package in Python [27]. Confidence intervals for both parameters are estimated by performing 10,000 bootstrap samples. For the zero-inflated Poisson model, the MLEs of *λ* and *σ* are calculated using scipy.optimize, with the confidence intervals estimated using a grid calculation. In this case *λ* is not the mean of the overall distribution but the mean of the Poisson component of the distribution. For the beta-Poisson distribution, 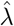 is given by the sample mean. In the Methods section we demonstrate that the contact parameter *N* has to be at least as large as *λ* for the beta-Poisson distribution to be well-defined. Based on this restriction and the fact that the model is valid when *N* = ∞, we define *ν* = *N* ^−1^ so that we can fit *ν* over the interval [0, 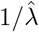). MLEs of Φ and *ν* are calculated using scipy.optimize. Confidence intervals on all three parameters are calculated by performing 10, 000 bootstrap samples. For each bootstrap sample we also calculate the probability of a case generating zero cases, the overdispersion, and the probability of generating a number of offspring in the upper 99^th^ percentile of the fitted Poisson distribution (i.e. the proportion of cases who are superspreaders according to the definition used by Lloyd-Smith *et al*. [2]) under each fitted distribution.

We fit each candidate model to eight sets of reconstructed secondary case data. Each of the datasets lists the number of cases in an outbreak which are inferred to have generated *x* secondary cases, for *x* running from zero up to the maximum number of secondary cases to be traced back to any one individual. Pneumonic plague, mpox (also known as monkeypox), and norovirus are each represented by single datasets. There are two sets of data from Ebola outbreaks and three from outbreaks of infections caused by novel coronaviruses: two of MERS and one of SARS. These datasets include examples of several different transmission routes: pneumonic plague and the two coronaviruses are spread through airborne transmission, norovirus mostly transmits through fecal-oral contact, and both Ebola and mpox are able to spread both through airborne transmission and through contact with bodily fluids (which itself includes fecal-oral contact).

The plague data is taken from a 2004 paper on pneumonic plague by Gani and Leach and combines data from several outbreaks [28]. This mixture of sources is clearly problematic since it can obscure differences between outbreaks caused by antigenic shift or simply socioeconomic disparities between the underlying populations (the data stretches across three continents and ninety years). Gani and Leach themselves find that a geometric distribution gives a good fit to the data, suggesting comparatively homogeneous spreading behaviour despite the range of sources. The mpox data is taken from a 1987 paper by Jezek *et al*. which reported on a mpox surveillance programme in what is now the Democratic Republic of Congo [13]. The first Ebola dataset is from a paper by Fasina et al. which constructs the transmission tree of a local outbreak in Nigeria during 2014 [29]. This outbreak was initiated by a single hospitalised patient to whom twelve subsequent cases were traced, suggesting a possible superspreading event. The other Ebola dataset is from a paper by Faye et al. which inferred transmission chains from line list data from the 2014 Ebola outbreak [30]. The line list data included all of the 193 confirmed probable and confirmed cases of Ebola in Guinea up to the time of the study, with 79% of these cases being assigned to transmission chains. The data is thus incomplete, but still provides us with a large sample of transmission behaviour in the epidemic. The SARS dataset is based on a transmission tree constructed by the Centre for Disease Control based on data from a SARS outbreak in Singapore, and contains clear evidence of superspreading behaviour [31]. The first of the two MERS-CoV datasets is from an outbreak of MERS which took place across three hospitals in South Korea in 2015, with each within-hospital outbreak initiated by the same index case who was moved between hospitals [32]. The data is extremely overdispersed, with most cases producing no subsequent cases but the index case producing over 80 subsequent cases. The other MERS-CoV dataset is from an outbreak in Saudi Arabia and is significantly less overdispersed [18]. The norovirus dataset was derived from a transmission tree constructed in a paper by Heijne et al. based on an outbreak in a psychiatric hospital in the Netherlands [33]. This dataset has a sub-geometric level of overdispersion.

The eight datasets are listed in Table 1; secondary case numbers which did not appear in any of the datasets are not listed. Each column has a bold entry marking the first element to appear after the 99^th^ percentile of the Poisson distribution with mean *R*, where *R* is the mean of the corresponding dataset, so that that entry (if nonzero) and any below it record the number of superspreading events. All of the datasets contain at least one superspreading event by this definition.

**Table 1.** Frequency of secondary case numbers by dataset. Entries in bold denote the superspreading boundary in each dataset.

| Secondary cases | Plague | Mpox | Ebola, Nigeria 2014 | Ebola, Guinea 2014 | SARS, Singapore 2003 | MERS, South Korea 2015 | MERS, Saudi Arabia 2015 | Norovirus, Netherlands 2012 |
| --- | --- | --- | --- | --- | --- | --- | --- | --- |
| 0 | 16 | 163 | 15 | 109 | 162 | 146 | 13 | 22 |
| 1 | 10 | 32 | 2 | 16 | 19 | 10 | 5 | 13 |
| 2 | 7 | **10** | 1 | 9 | 8 | 4 | 4 | 6 |
| 3 | 2 | 2 | 1 | 5 | **7** | 1 | 1 | 3 |
| 4 | 3 | 0 | **0** | **5** | 0 | **0** | **0** | **1** |
| 5 | **1** | 1 | 0 | 2 | 0 | 1 | 0 | 1 |
| 6 | 1 | 0 | 0 | 0 | 0 | 1 | 0 | 0 |
| 7 | 0 | 0 | 0 | 0 | 1 | 0 | 1 | 0 |
| 8 | 0 | 0 | 0 | 1 | 0 | 0 | 0 | 0 |
| 9 | 0 | 0 | 0 | 3 | 0 | 0 | 0 | 0 |
| 12 | 0 | 0 | 1 | 0 | 1 | 0 | 0 | 0 |
| 14 | 0 | 0 | 0 | 1 | 0 | 0 | 0 | 0 |
| 17 | 0 | 0 | 0 | 1 | 0 | 0 | 0 | 0 |
| 21 | 0 | 0 | 0 | 0 | 1 | 0 | 0 | 0 |
| 23 | 0 | 0 | 0 | 0 | 1 | 1 | 0 | 0 |
| 38 | 0 | 0 | 0 | 0 | 0 | 1 | 0 | 0 |
| 40 | 0 | 0 | 0 | 0 | 1 | 0 | 0 | 0 |
| 81 | 0 | 0 | 0 | 0 | 0 | 1 | 0 | 0 |
| Total | 53 | 63 | 19 | 145 | 159 | 174 | 23 | 43 |

The MLEs of the beta-Poisson model parameters fitted to each dataset are listed in Table 3 and plotted in Fig 2. MLEs and confidence intervals for the negative binomial and ZIP models are provided for reference in Section 7 of S1 Appendix. The AIC achieved by each model fitted to each dataset is listed in Table 2, and the log likelihood ratios obtained by comparing the maximum likelihood attained by the beta-Poisson to those obtained by each of the other models are listed in Table 4. In Section 8 of Appendix S1 we perform a sensitivity analysis on the beta-Poisson model by exploring the likelihood surface of the model parameters around each MLE. As a generalisation of all the other models under consideration, the beta-Poisson will always be able to obtain the highest likelihood; if another distribution appeared to offer a higher maximum likelihood, we could simply parameterise the beta-Poisson to match that specific case. Despite this, Table 2 reveals that is never the optimal choice in terms of AIC, with the improvement in likelihood never substantial to justify the addition of an extra parameter relative to the negative binomial distribution. For secondary case distributions with low levels of overdispersion (plague, mpox, norovirus, MERS in Saudi Arabia, see Table 5 and Fig 4), the geometric distribution attains the smallest AIC, although in all of these cases a comparison between the geometric and beta-Poisson models using the likelihood ratio test at any reasonable confidence level will lead us to reject the geometric model. In the other four cases (the two Ebola datasets, SARS, and MERS in South Korea), the negative binomial model is optimal in terms of AIC, and the likelihood ratio between the beta-Poisson and negative binomial models is close to 1. In the case of SARS and the South Korean MERS outbreak, the beta-Poisson model fits very decisively to its negative binomial extreme. Both these datasets include one or more dramatic superspreading events where a single individual is responsible for a substantial proportion of the observed cases, and in these cases the homogeneous contact behaviour assumed by the beta-Poisson model away from its negative binomial limit will struggle to capture this type of event, which require that the individual responsible have an unusually high number of social contacts during their infectious period.

**Table 2.** Akaike information criterion at MLE by model and dataset. The underlined entry of each row is the minimal value of AIC attained for that dataset.

| Dataset | Poisson | Geometric | Neg. bin. | ZIP | Beta-Poisson |
| --- | --- | --- | --- | --- | --- |
| Plague | 136.84 | <u>129.1</u> | 130.62 | 131.89 | 132.24 |
| Mpox | 309.1 | <u>295.9</u> | 296.63 | 298.02 | 298.63 |
| Ebola, Nigeria 2014 | 86.89 | 56.04 | <u>47.45</u> | 58.47 | 49.41 |
| Ebola, Guinea 2014 | 602.41 | 413.56 | <u>358.39</u> | 416.11 | 360.37 |
| SARS, Singapore 2003 | 902.35 | 496.15 | <u>357.47</u> | 579.32 | 359.47 |
| MERS, South Korea 2015 | 1230.77 | 473.15 | <u>224.66</u> | 618.44 | 226.66 |
| MERS, Saudi Arabia 2015 | 76.14 | <u>67.13</u> | 68.89 | 71.84 | 70.89 |
| Norovirus, Netherlands 2012 | 128.8 | <u>125.28</u> | 126.57 | 127.25 | 128.46 |

**Table 3.** Maximum likelihood estimates of beta-Poisson model parameters by dataset, 95% confidence intervals in parentheses.

| Dataset | $\lambda$ | $\Phi$ | $\nu$ |
| --- | --- | --- | --- |
| Plague | 1.32 (0.88, 1.82) | 0.58 (0.0, 7.88) | 0.25 (0.0, 0.51) |
| Mpox | 0.3 (0.22, 0.4) | 1.93 (0.0, 9.31) | 0.0 (0.0, 1.68) |
| Ebola, Nigeria 2014 | 0.95 (0.15, 2.3) | 0.12 (0.0, 10.22) | 0.05 (0.0, 6.67) |
| Ebola, Guinea 2014 | 0.95 (0.61, 1.36) | 0.18 (0.1, 0.34) | 0.01 (0.0, 0.14) |
| SARS, Singapore 2003 | 0.79 (0.36, 1.38) | 0.12 (0.05, 0.42) | 0.0 (0.0, 0.0) |
| MERS, South Korea 2015 | 1.05 (0.2, 2.31) | 0.04 (0.01, 0.4) | 0.0 (0.0, 0.02) |
| MERS, Saudi Arabia 2015 | 0.96 (0.42, 1.62) | 0.75 (0.0, 12.41) | 0.0 (0.0, 1.64) |
| Norovirus, Netherlands 2012 | 0.93 (0.61, 1.28) | 1.07 (0.0, 16.52) | 0.31 (0.0, 1.18) |

**Table 4.**
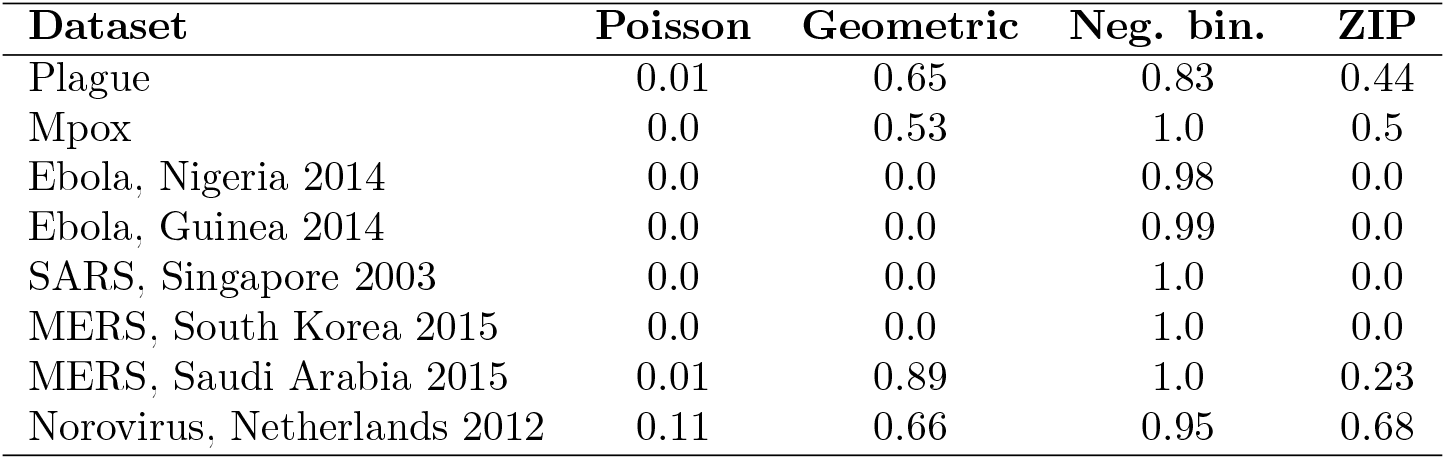
Likelihood ratios of beta-Poisson to negative binomial and ZIP models under each dataset.

**Table 5.** Overdispersion of each maximum likelihood distribution, 95% confidence intervals in parentheses.

| Dataset | Observed | Geometric | Neg. bin. | ZIP | Beta-Poisson |
| --- | --- | --- | --- | --- | --- |
| Plague | 0.79 | 1.32 (0.88, 1.83) | 0.92 (0.12, 1.98) | 0.54 (0.06, 1.1) | 0.81 (0.12, 1.46) |
| Mpox | 0.52 | 0.3 (0.22, 0.4) | 0.52 (0.1, 1.05) | 0.41 (0.11, 0.76) | 0.52 (0.1, 0.99) |
| Ebola, Nigeria 2014 | 6.42 | 0.95 (0.15, 2.3) | 6.89 (0.0, 24.55) | 2.76 (0.0, 7.08) | 5.64 (0.0, 9.78) |
| Ebola, Guinea 2014 | 5.07 | 0.95 (0.61, 1.36) | 5.26 (2.66, 8.77) | 2.29 (1.44, 3.25) | 5.0 (2.3, 8.21) |
| SARS, Singapore 2003 | 16.3 | 0.79 (0.36, 1.38) | 8.5 (2.07, 18.87) | 3.21 (1.25, 5.51) | 8.5 (2.01, 18.47) |
| MERS, South Korea 2015 | 47.55 | 1.05 (0.2, 2.31) | 28.18 (1.91, 77.98) | 7.65 (1.24, 16.54) | 28.18 (1.77, 68.3) |
| MERS, Saudi Arabia 2015 | 1.48 | 0.96 (0.42, 1.67) | 1.33 (0.0, 4.0) | 0.76 (0.0, 1.82) | 1.33 (0.0, 3.05) |
| Norovirus, Netherlands 2012 | 0.51 | 0.93 (0.61, 1.3) | 0.56 (0.0, 1.39) | 0.37 (0.0, 0.86) | 0.52 (0.0, 1.11) |

**Fig 2.**
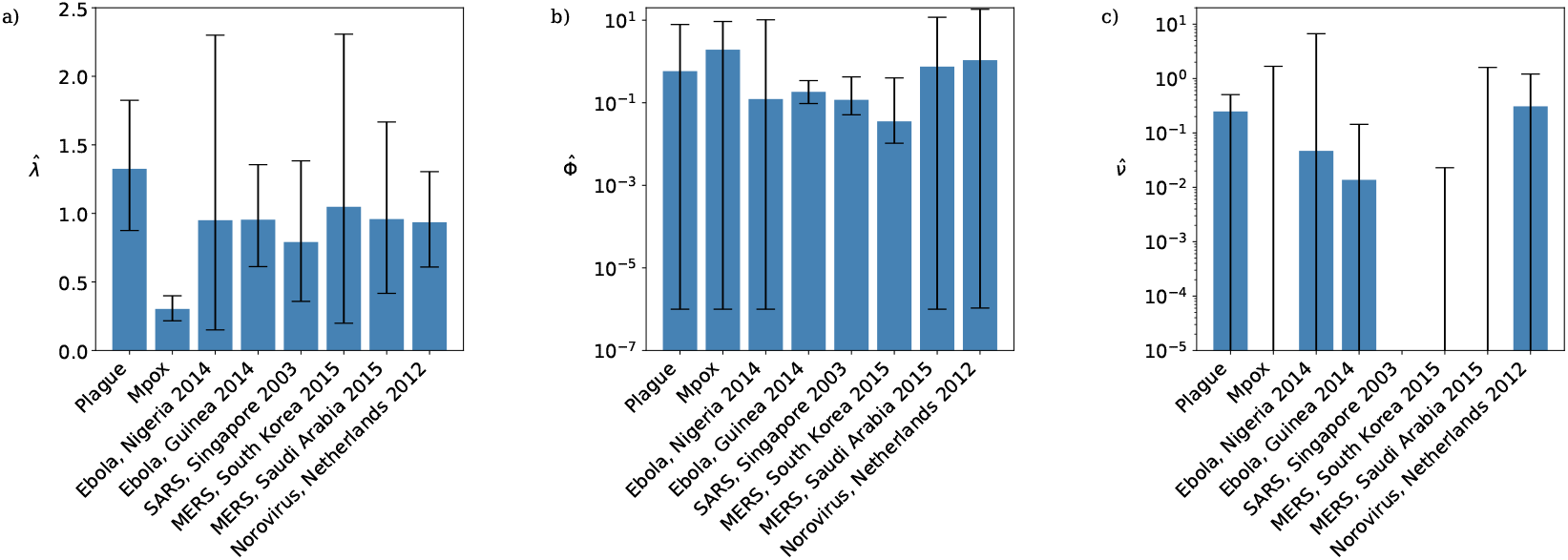
Maximum likelihood estimates of the beta-Poisson model parameters by dataset: a) basic reproductive ratio *λ*; b) overdispersion parameter Φ; c) inverse contact parameter *ν*. Black lines are 95% confidence intervals. In plot c) MLEs and lower confidence bounds of 0 are not shown.

The lower portions of the empirical secondary case distributions and model distributions with fitted MLE parameters are plotted in Fig 3. For the mpox data, Singapore SARS data, and both sets of MERS data the maximum likelihood distribution is identical for the negative binomial and beta-Poisson distributions, and so only the fitted negative binomial is shown. In those cases where the beta-Poisson does not fit to its negative binomial limit, the fitted beta-Poisson distribution does not appear to differ dramatically from the fitted negative binomial in its description of the transmission potential of low-progeny cases. Figure 4 and Table 5 suggests that where it fits to a distinct distribution the beta-Poisson performs similarly to the negative binomial in its ability to capture the level of overdispersion in the data, although with slightly narrower 95% confidence intervals. The overdispersion in the secondary case datasets is driven both by superspreading events and by an abundance of cases producing zero secondary cases. Figure 5 and Table 6 reveal that the beta-Poisson and negative binomial models both predict similar proportions of superspreaders when fitted to each dataset, and that in general these proportions are not necessarily close to those observed in the data. Figure 6 and Table 7 demonstrate that both of these models fit closely to the observed proportion of zero-progeny cases, as does the ZIP distribution.

**Fig 3.**
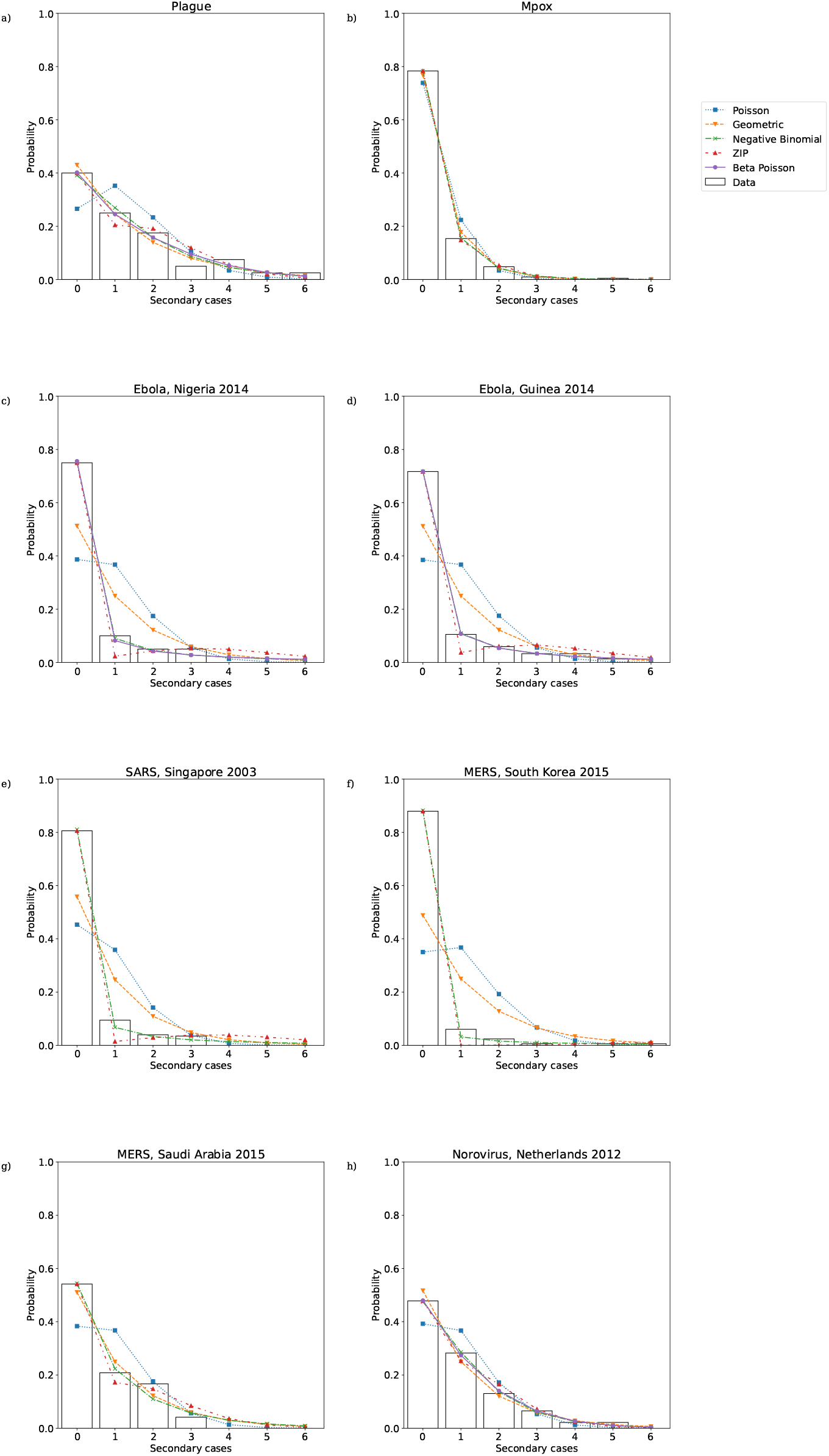
Lower portion of maximum likelihood offspring distributions fitted to secondary case data from a) plague; b) Mpox; c) Ebola, Nigeria 2014; d) Ebola, Guinea 2014; e) SARS, Singapore 2003; f) MERS, South Korea 2015; g) MERS, Saudi Arabia 2015; h) Norovirus, Netherlands 2012.

**Fig 4.**
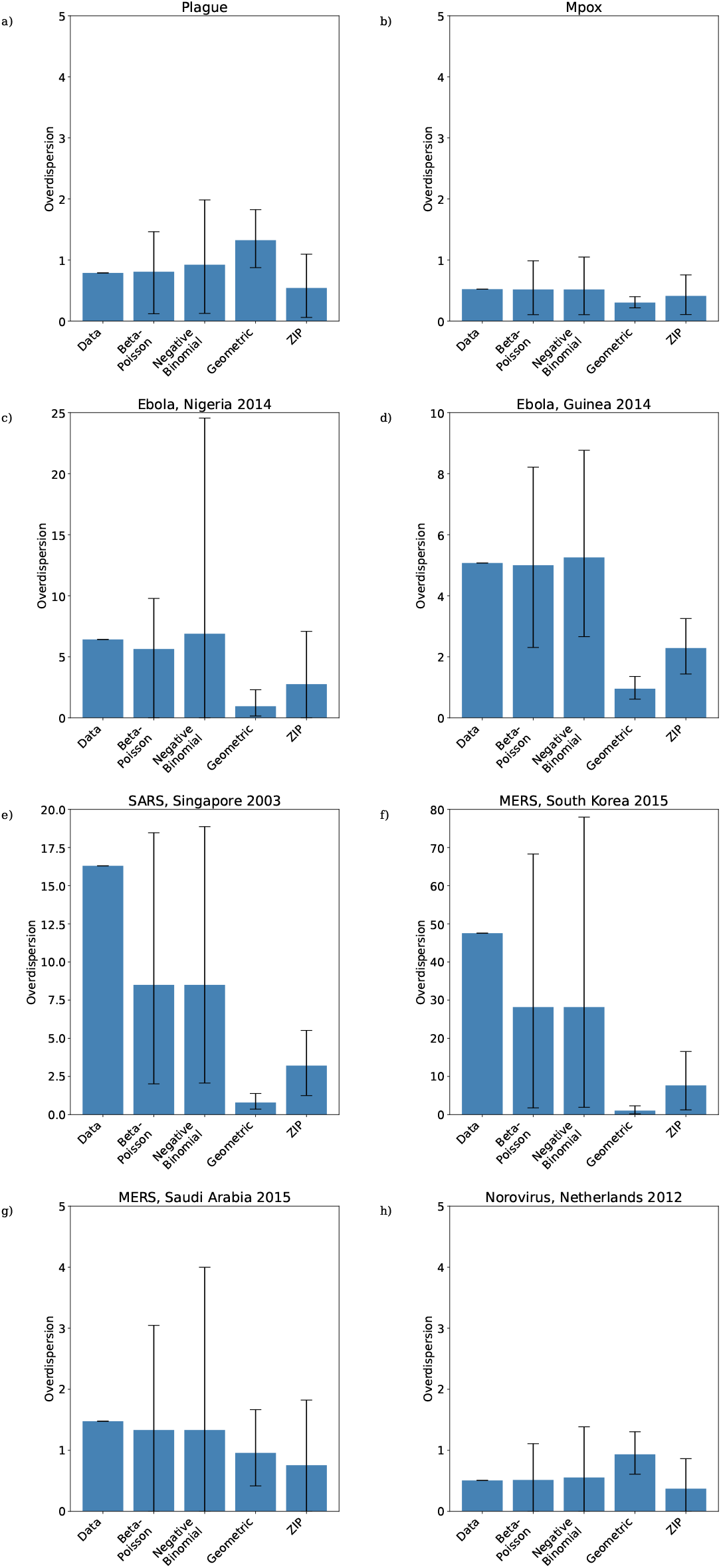
Overdisperion of maximum likelihood offspring distributions fitted to reconstructed transmission trees from a) plague; b) Mpox; c) Ebola, Nigeria 2014; d) Ebola, Guinea 2014; e) SARS, Singapore 2003; f) MERS, South Korea 2015; g) MERS, Saudi Arabia 2015; h) Norovirus, Netherlands 2012. Black lines are 95% confidence intervals.

**Fig 5.**
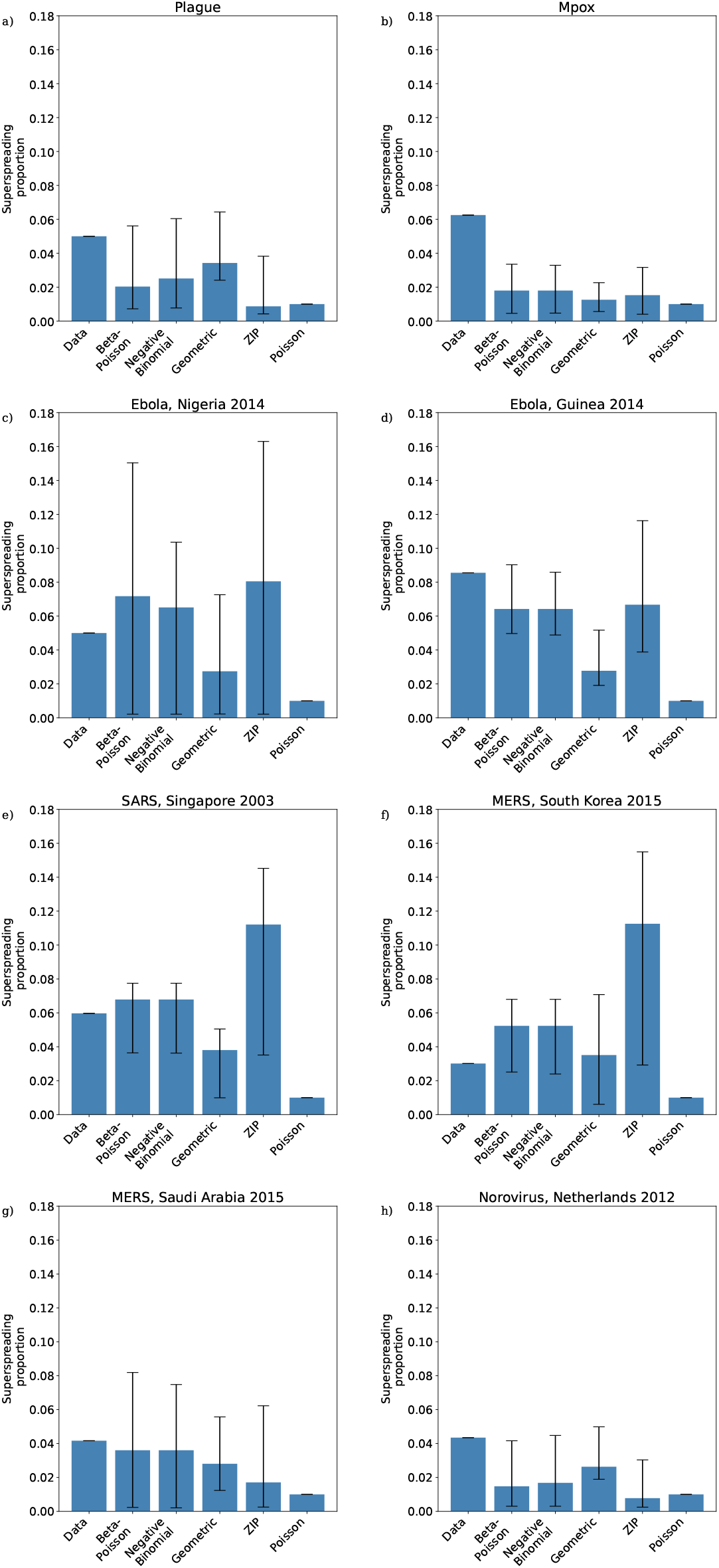
Proportion of superspreaders in maximum likelihood offspring distributions fitted to reconstructed transmission trees from a) plague; b) Mpox; c) Ebola, Nigeria 2014; d) Ebola, Guinea 2014; e) SARS, Singapore 2003; f) MERS, South Korea 2015; g) MERS, Saudi Arabia 2015; h) Norovirus, Netherlands 2012. Black lines are 95% confidence intervals.

**Fig 6.**
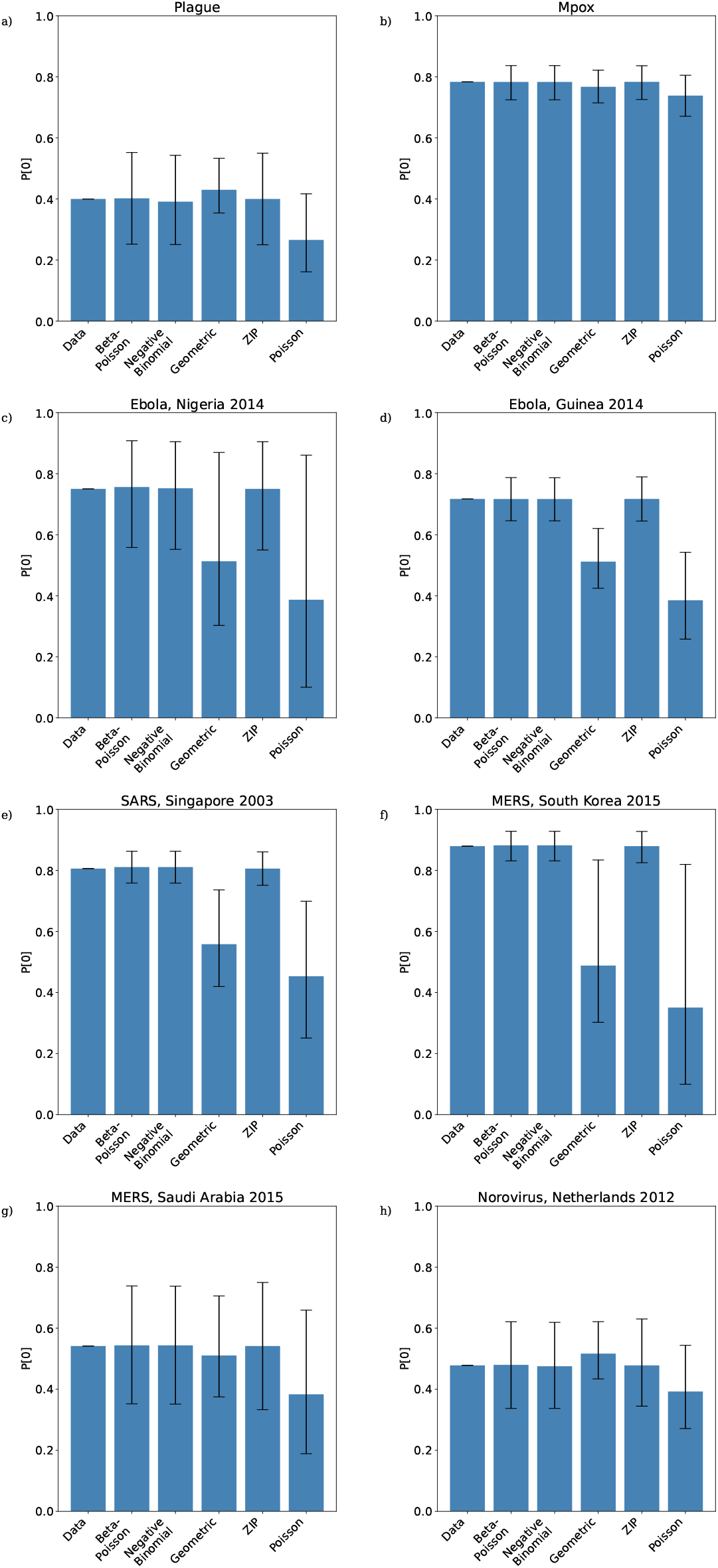
Proportion of zeros in maximum likelihood offspring distributions fitted to reconstructed transmission trees from a) plague; b) Mpox; c) Ebola, Nigeria 2014; d) Ebola, Guinea 2014; e) SARS, Singapore 2003; f) MERS, South Korea 2015; g) MERS, Saudi Arabia 2015; h) Norovirus, Netherlands 2012. Black lines are 95% confidence intervals.

**Table 6.**
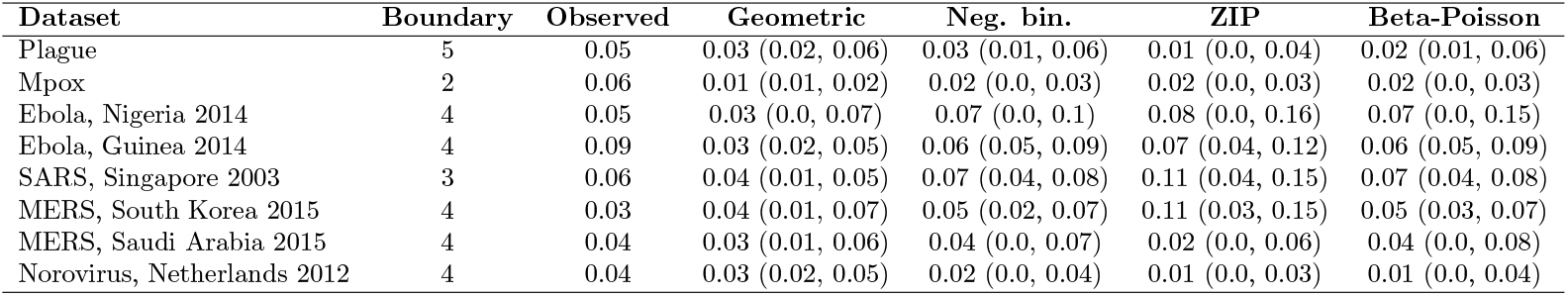
Superspreading boundary (99th percentile of fitted Poisson distribution) and proportion of cases above this boundary for each maximum likelihood distribution, 95% confidence intervals in parentheses.

| Dataset | Boundary | Observed | Geometric | Neg. bin. | ZIP | Beta-Poisson |
| --- | --- | --- | --- | --- | --- | --- |
| Plague | 5 | 0.05 | 0.03 (0.02, 0.06) | 0.03 (0.01, 0.06) | 0.01 (0.0, 0.04) | 0.02 (0.01, 0.06) |
| Mpox | 2 | 0.06 | 0.01 (0.01, 0.02) | 0.02 (0.0, 0.03) | 0.02 (0.0, 0.03) | 0.02 (0.0, 0.03) |
| Ebola, Nigeria 2014 | 4 | 0.05 | 0.03 (0.0, 0.07) | 0.07 (0.0, 0.1) | 0.08 (0.0, 0.16) | 0.07 (0.0, 0.15) |
| Ebola, Guinea 2014 | 4 | 0.09 | 0.03 (0.02, 0.05) | 0.06 (0.05, 0.09) | 0.07 (0.04, 0.12) | 0.06 (0.05, 0.09) |
| SARS, Singapore 2003 | 3 | 0.06 | 0.04 (0.01, 0.05) | 0.07 (0.04, 0.08) | 0.11 (0.04, 0.15) | 0.07 (0.04, 0.08) |
| MERS, South Korea 2015 | 4 | 0.03 | 0.04 (0.01, 0.07) | 0.05 (0.02, 0.07) | 0.11 (0.03, 0.15) | 0.05 (0.03, 0.07) |
| MERS, Saudi Arabia 2015 | 4 | 0.04 | 0.03 (0.01, 0.06) | 0.04 (0.0, 0.07) | 0.02 (0.0, 0.06) | 0.04 (0.0, 0.08) |
| Norovirus, Netherlands 2012 | 4 | 0.04 | 0.03 (0.02, 0.05) | 0.02 (0.0, 0.04) | 0.01 (0.0, 0.03) | 0.01 (0.0, 0.04) |

**Table 7.** Probabliity of a case generating zero secondary cases under each maximum likelihood distribution, 95% confidence intervals in parentheses.

| Dataset | Poisson | Observed | Geometric | Neg. bin. | ZIP | Beta-Poisson |
| --- | --- | --- | --- | --- | --- | --- |
| Plague | 0.4 | 0.27 (0.16, 0.42) | 0.43 (0.35, 0.53) | 0.39 (0.25, 0.54) | 0.4 (0.25, 0.55) | 0.4 (0.25, 0.55) |
| Mpox | 0.78 | 0.74 (0.67, 0.81) | 0.77 (0.71, 0.82) | 0.78 (0.72, 0.84) | 0.78 (0.73, 0.84) | 0.78 (0.72, 0.84) |
| Ebola, Nigeria 2014 | 0.75 | 0.39 (0.1, 0.86) | 0.51 (0.3, 0.87) | 0.75 (0.55, 0.9) | 0.75 (0.55, 0.9) | 0.76 (0.56, 0.91) |
| Ebola, Guinea 2014 | 0.72 | 0.39 (0.26, 0.54) | 0.51 (0.42, 0.62) | 0.72 (0.65, 0.79) | 0.72 (0.64, 0.79) | 0.72 (0.65, 0.79) |
| SARS, Singapore 2003 | 0.81 | 0.45 (0.25, 0.7) | 0.56 (0.42, 0.74) | 0.81 (0.76, 0.86) | 0.81 (0.75, 0.86) | 0.81 (0.76, 0.86) |
| MERS, South Korea 2015 | 0.88 | 0.35 (0.1, 0.82) | 0.49 (0.3, 0.83) | 0.88 (0.83, 0.93) | 0.88 (0.83, 0.93) | 0.88 (0.83, 0.93) |
| MERS, Saudi Arabia 2015 | 0.54 | 0.38 (0.19, 0.66) | 0.51 (0.37, 0.71) | 0.54 (0.35, 0.74) | 0.54 (0.33, 0.75) | 0.54 (0.35, 0.74) |
| Norovirus, Netherlands 2012 | 0.48 | 0.39 (0.27, 0.54) | 0.52 (0.43, 0.62) | 0.48 (0.34, 0.62) | 0.48 (0.34, 0.63) | 0.48 (0.34, 0.62) |

## Discussion

The beta-Poisson distribution will by definition consistently achieve a closer fit to transmission chain data than the more commonly used distributions which we have considered here, but our results demonstrate these models outperform it equally consistently on grounds of parsimony. This suggests that the topology of transmission chains is simple enough to be summarised in just two parameters. For transmission chains with low levels of overdispersion the parameter Φ appears to be unidentifiable, whereas when overdispersion is high a good fit can be achieved using a negative binomial model. Although the contact rates fitted in the two Ebola examples were large but clearly not infinite, the negative binomial model still outperformed the beta-Poisson in terms of AIC. Despite this lack of improvement over the negative binomial, the explicit contact parameter in the beta-Poisson model introduces a capacity for explicitly modelling non-pharmaceutical interventions which act to reduce the number of contacts during an individual’s infectious period. A limitation of the beta-Poisson model is that this number of contacts is limited to Poisson behaviour. In reality it may be the social variation in number of contacts (suggesting a Negative binomial distribution on number of contacts) and this may be a natural additional model complexity to consider in future research. However, the model fitting shown here suggests that additional parametric structure is unlikely to be supported by available data. Like the negative binomial distribution, the beta-Poisson distribution is able to capture the high proportion of cases in infectious disease outbreaks which do not generate any secondary cases, but it does not appear to offer a substantial improvement in the ability to quantify the superspreading events which characterise many outbreaks.

In the Supplementary Information we demonstrate that the beta-Poisson model’s outbreak size probability formula is extremely cumbersome. In the absence of detailed transmission chain data branching process models are often parameterised using the total number of cases in an epidemic or set of epidemics [2, 14], and this cumbersome formula makes the relevant likelihood calculations substantially more computationally intensive than those associated with other commonly used models. The presence of an explicit contact parameter also introduces complexities in dealing with outbreak size data, since there is no a priori reason to think that this *N* will stay fixed across disparate population settings. This means that any dataset which include the sizes of outbreaks in multiple distinct settings will potentially include samples from distributions with different underlying parameters, which is particularly problematic in situations where data is combined from multiple countries and over long time intervals.

Although the limitations introduced by population specific parameters are obvious in the beta-Poisson model, the same concerns are relevant to any branching process model, and indeed to many other classes of epidemiological model. In the Poisson, geometric, and negative binomial models, the parameters *λ* and *θ* encode a combination of physiological and social factors. Whereas we expect physiological factors to be broadly disease specific, these social factors are by no means consistent from population to population. This means that the same care needs to be taken in using outbreak size data to parameterise these models as is required for the beta-Poisson model. Population-specific parameters also need to be taken into account when simulating outbreaks. This has obvious repercussions in modelling infections on an international basis, with pandemic diseases by definition moving across population boundaries. Slightly less obvious is the implications for using data from very specific and unrepresentative social environments such as hospitals. This is a particularly important consideration since transmission chain data often comes from these types of settings. Of the example datasets we studied here, the Nigerian Ebola [29], South Korean MERS [18], Saudi Arabian MERS, and Dutch norovirus [33] datasets all come from sources which mention hospital-based transmission. Although our focus here has been on branching process models, compartmental models also include population-specific parameters, with the transmission rate in the standard SIR model subsuming physiological processes and contact behaviours into a single parameter. In fact, the basic reproductive ratio *R*_0_, while often quoted as a disease-specific quantity, is also population-specific. Even though this is not a particularly novel point (the tables of basic reproductive ratios in Anderson and May’s textbook is careful to list locations along with pathogens [34]), it is still an important one which is highlighted by the beta-Poisson model’s explicit inclusion of a social contact parameter.

Thinking carefully about population-specific and disease-specific parameters leads us to the natural question of whether, in its (*α*_1_, *α*_2_, *N*) formulation, the beta-Poisson model actually disentangles its underlying social and physiological processes. Such a decoupling would allow us to fit the parameters to outbreak data from a given population setting and then simulate outbreaks in different populations by changing *N* whilst keeping *α*_1_ and *α*_2_ constant. At first glance, one might consider it valid to label *N* a social parameter and *α*_1_ and *α*_2_ physiological parameters, with the beta-distributed transmission probability *p* a function of a given case’s physiological response to the infection. However, this transmission probability and the distribution is is drawn from can potentially encode information about contact behaviour as well as physiological shedding behaviour. For instance, while children are known to shed greater quantities of influenza virus than adults [35], contact survey also suggests that a higher proportion of their contacts are physical than those of adults [25], with both of these factors being relevant to transmission probabilities. This suggests that although the Poisson parameter *N* relates only to social behaviours, the beta distributed transmission probability incorporates both physiological and social factors. In fact, even if the beta distribution only summarised the physiological responses of a given population, any beta distribution parameterised to outbreak data is still specific to the outbreak’s underlying population, which may be an unrepresentative subset of some larger population of interest. To see why this is the case, consider a dataset drawn from an outbreak confined to a single hospital ward. Since children and adults are usually assigned to separate wards, a beta-Poisson distribution parameterised using data from an adult ward will contain no information about the shedding behaviour of children, which as we have mentioned already can differ from that of adults. For example, the South Korean MERS dataset, collated from three hospital wards, contains only one case in a person under 18 [32]. As with the considerations surrounding the social contact parameter, this problem is clearly present in other types of model, but is made more obvious by the mechanistic structure of our model. The idea that outbreak data can be specific to an underlying population suggests that some conception of “representativeness” needs to be taken into account when making predictions using parameterised models.

## Methods

The model description and log-likelihood formula in this section is sufficient to reproduce the results presented in this paper. To make our account of the beta-Poisson model more complete, in Appendix S1 we outline moment calculations (Section 1), the probability generating function (Section 2), the branching process extinction probability (Section 3), the minor outbreak size distribution (Section 4), the distribution of attack rates in a major outbreak (Section 5), and likelihood calculations using outbreak size data (Section 6).

### Model description

The beta-Poisson model describes the person-to-person spread of a pathogen in a population where contact behaviour is homogeneous but transmission behaviour varies from person to person. During their infectious period an infectious individual contacts at the points of a Poisson process, so that their total contacts during this time follow Poisson distribution with mean *N*. For each case a transmission probability *p* is chosen from a beta distribution with parameters (*α*_1_, *α*_2_), and each of their contacts results in a new infection with this probability. This variation in infection probability captures an individual-level variability in infectiousness, which has been suggested as a possible driver of superspreading in Ebola [15]. Given *p*, the transmission process is a Poisson process with intensity *pN*, and so the total number of infections generated during an individual’s infectious period is Poisson distributed with mean *pN*. With this in mind, we can integrate with respect to *p* to obtain the probability density function of the beta-Poisson distribution:

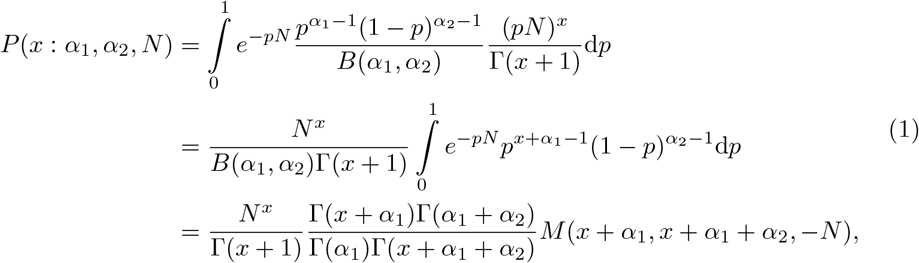

where *M* (*a, b, x*) is the confluent hypergeometric function. We will use several properties of this function when required but will not discuss its properties in detail. For more information see, for example, Abramowitz and Stegun [36]. Using the identity

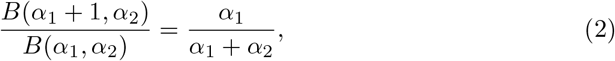

the mean *λ* of the distribution is given by

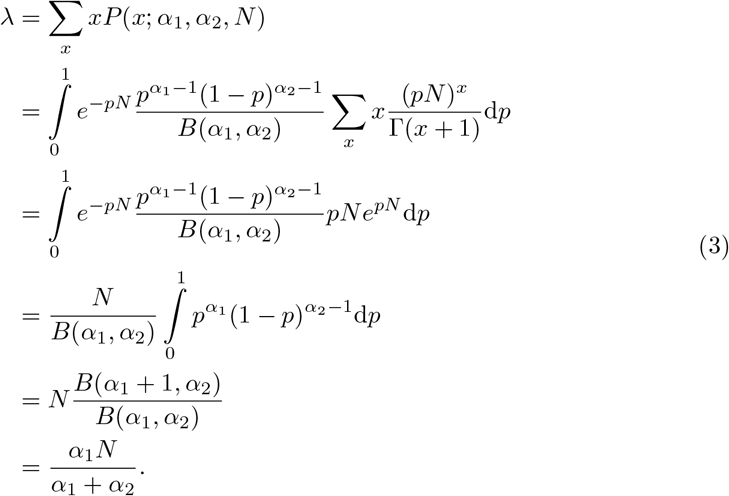

From this is is immediately clear that *λ* ≤ *N*. The notation *λ* is chosen by analogy with the standard notation for the mean of the (unmixed) Poisson distribution, and in our epidemiological application *λ* is the basic or effective reproductive ratio (depending on context). The beta distribution parameters *α*_1_ and *α*_2_ lack an intuitive interpretation in terms of transmission behaviour, but by making the substitution 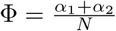 we can express the distribution in terms of *λ*, Φ, and *N* :

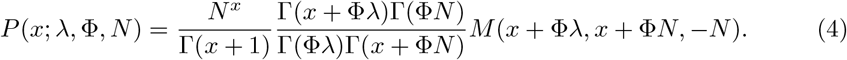

When its first two arguments are equal, the confluent hypergeometric function is given by the exponential of its third argument, and so the beta-Poisson distribution reduces to the (unmixed) Poisson distribution when *λ* = *N*.

In the limit *N* ⟶ ∞, the beta-Poisson distribution is equivalent to the negative binomial distribution with parameters *λ* and *θ* = Φ^−1^. To see this, we first note that

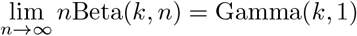

and that

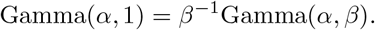

From the definition of the beta-Poisson distribution, a beta-Poisson random variable is drawn from a Poisson distribution with mean *Np* where *p* ∼ Beta(*λ*Φ, (*N* − *λ*)Φ). From the identities stated above it follows that

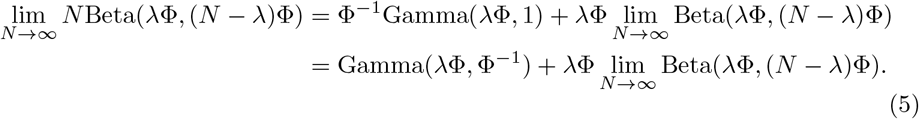

It follows that the beta-Poisson distribution can be expressed as the sum of a Gamma-Poisson mixture with Gamma parameters 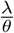 and *θ* = Φ^−1^ and a beta-Poisson mixture with parameters *λ*Φ and (*N* − *λ*)Φ, with the latter term scaled by a factor of *λ*Φ. This Poisson-beta mixture is slightly different to the beta-Poisson model we are considering in this study, since our model applies a scaling of *N* to the beta-distributed random variable before feeding it into the Poisson distribution. The *r*th moment of a beta distribution with parameters (*α*_1_, *α*_2_) is given by the product

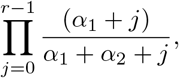

and so in the limit *N* ⟶ ∞, all of the moments of the distribution Beta(*λ*Φ, (*N* − *λ*)Φ) will tend to zero. In Section 1 of Appendix S1 we show that because the *r*th moment of a mixed Poisson distribution is given by a weighted sum of the first *r* moments of its mixing distribution [16], the moments of the beta-Poisson mixture will also tend to zero as *N* tends to infinity. Thus, as *N* ⟶ ∞, this beta-Poisson mixture will tend to a point mass at zero. This leaves us with only the Gamma-Poisson mixture term of our sum, which is precisely the negative binomial distribution with parameters (*λ, θ*).

The variance of the beta-Poisson distribution is given by

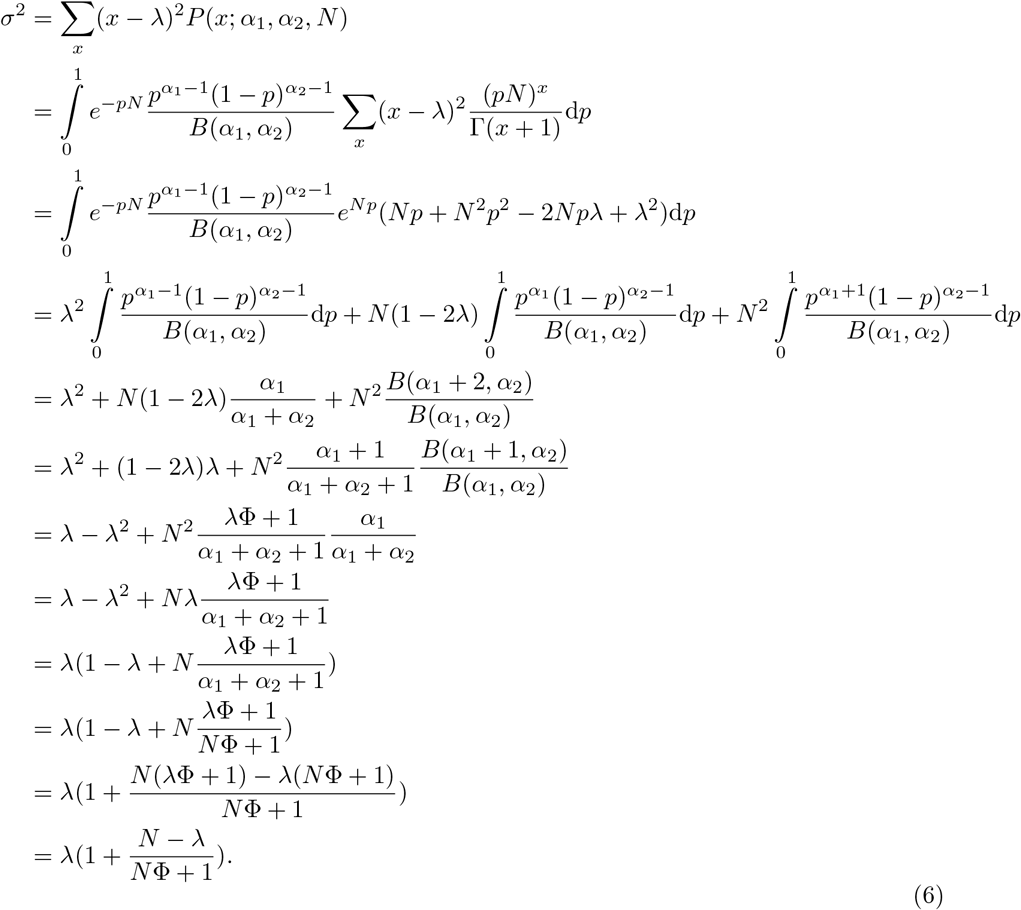

The variance-to-mean ratio *E* is given by

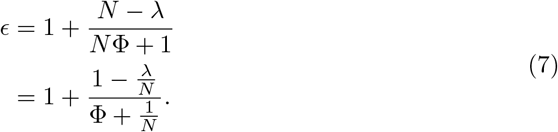

In the limit *N* ⟶ ∞, 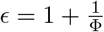, in keeping with our interpretation of Φ as the reciprocal of the overdispersion parameter *θ* from the negative binomial distribution. When *λ* = *N* the variance-to-mean ratio is 1, in keeping with our earlier statement that this is the (unmixed) Poisson distribution.

We can express the standard beta distribution parameters as follows:

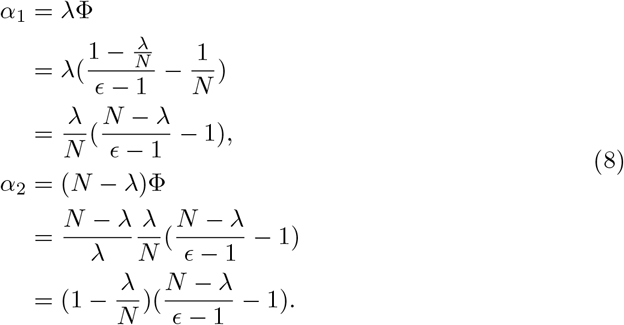

Since both these quantities are strictly positive and *N* ≥ *λ*, we see that *N* − *λ* ≥ *ϵ* − 1 ≥ 0. The condition that *ϵ* ≥ 1 means the distribution is overdispersed relative to the Poisson parameter except when *λ* = *N*, i.e. *α*_2_ = 0. In this scenario the beta mean is 1 and so all of the binomial transmission trials are successful, meaning our transmission distribution is precisely the Poisson distribution with mean *N*. We also have an upper bound on the variance, *σ*^2^ ≤ (1 + *N* − *λ*)*λ*.

### Likelihood calculations using secondary case data

The log-likelihood of parameters (*λ*, Φ, *N*) given the secondary case data (*x*_1_, …, *x*_*K*_) is given by

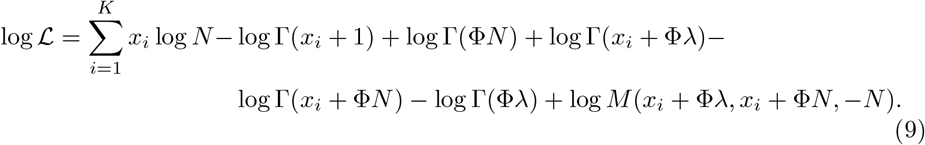

The confluent hypergeometric function is not differentiable with respect to its first two arguments, meaning we can not find maximum likelihood estimates analytically. However, the branching process structure of our model means that the MLE of *λ* is the sample mean of the secondary case data [11]. This reduces the calculation of an MLE to a two-dimensional problem which can be solved numerically.

## Data Availability

The code used in this study is available at https://github.com/JBHilton/beta-poisson-epidemics.

## Acknowledgments

The authors would like to thank Louise Dyson and Jonathan Read for their helpful comments on this work.

## Appendix

### 1 Moment calculations

The moments of the beta-Poisson distribution are relatively easy to calculate. For a mixed Poisson process with rate *Y* (where *Y* is some random variable), the *r*th moment 𝔼 [*X*^*r*^] is given by

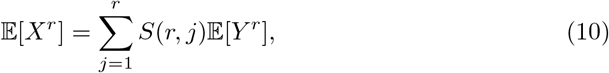

where *S*(*r, j*) denotes the Stirling numbers of the second kind [16]. In our model, *Y* = *Np*, so **E**[*Y* ^*r*^] = *N*^*r*^𝔼 [*p*^*r*^]. Using identity 3 from the main text, we find that

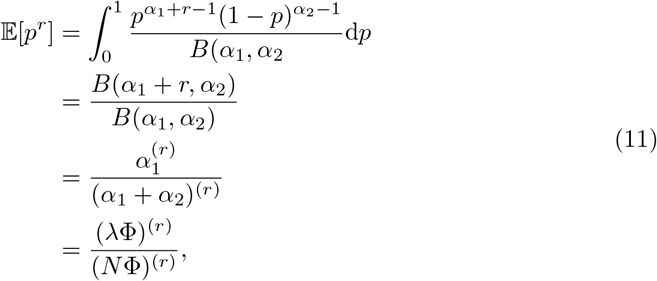

where we have applied Equation 3 from the main text repeatedly to write the ratio of beta functions in terms of the Pochhamer function defined by

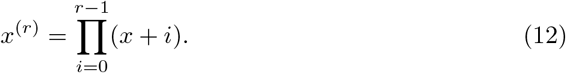

It follows that the *r*th moment of the beta-Poisson distribution is given by the formula

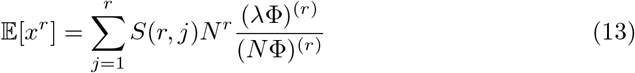

Using the fact that *S*(2, 1) = *S*(2, 2) = 1 (see, for example, Abramowitz and Stegun [36]), for *r* = 2 we find

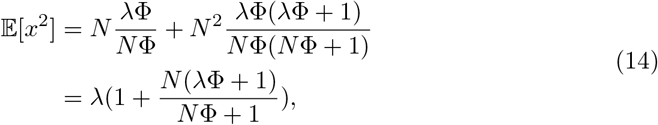

and so

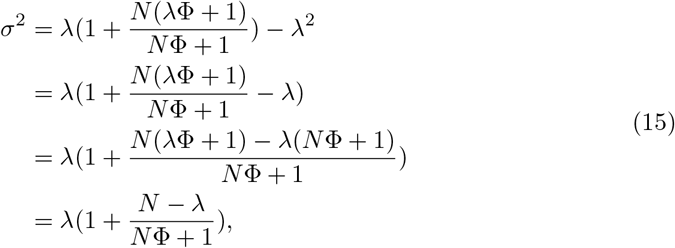

in agreement with the expression we found through direct calculation in the Methods section of the main text.

Alongside the mean and variance, another moment which may be of interest in the context of outbreak modelling is kurtosis. This can be interpreted as measuring a distribution’s “tailedness”, which is of interest since superspreading events by definition belong to the tail of the offspring distribution. The kurtosis is given by

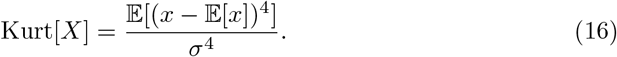

Using Equation 10, the calculation of kurtosis is as follows (we multiply by *σ*^4^ for notational convenience, since nothing particularly cancels out):

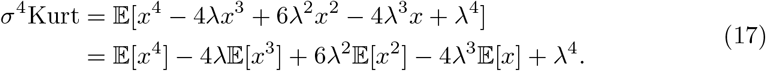

Using Equation 10 and substituting in the required Stirling numbers, we get the following four equations for the moments of *x*:

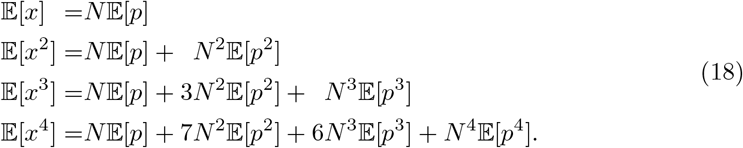

Substituting these expressions into Equation 17, we get

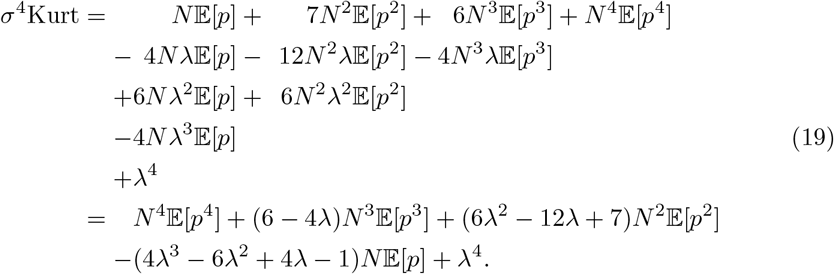

Since *N* 𝔼 [*p*] = *λ*, the last two terms add together to give

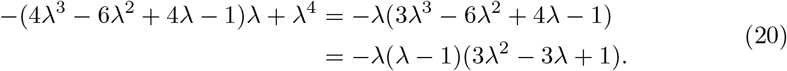

Substituting this and Equation 11 into Equation 19, we get

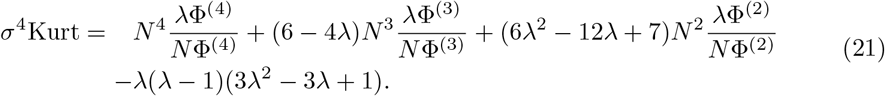

We leave the Pochhammer symbols as is, since the formula is unlikely to clean up any more than this. Since *σ*^2^ is linear in *λ*, it follows that the kurtosis will end up being quadratic in *λ*.

### 2 Generating function

The beta-Poisson distribution’s probability generating function is necessary for calculating the extinction probability and outbreak size distribution.

The generating function of a mixed Poisson distribution with mixing distribution pdf *g*(*λ*) is given by [16]

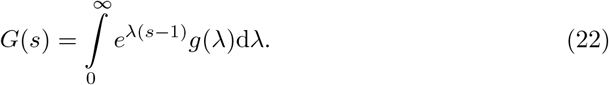

Thus, the pgf of a beta-Poisson distributed random variable *X* is

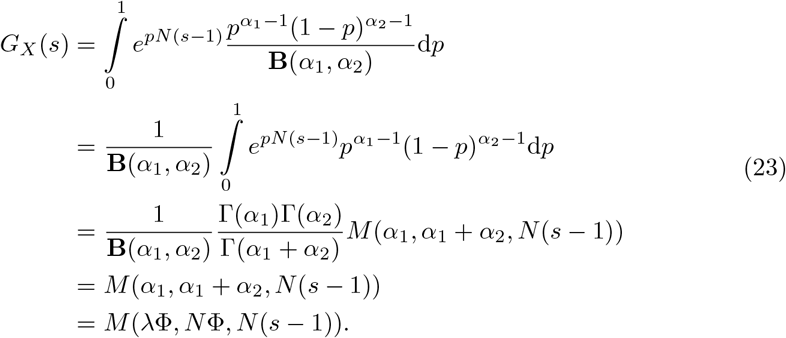

The generating function of the random variable *Z*, the total cases generated by *K* cases, is then

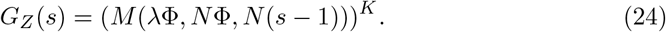

### 3 Extinction probability

Extinction occurs with probability one for branching process epidemics with *R*_0_ ≤ 1. The extinction probability of a branching process epidemic with *R*_0_ *>* 1 is given by the unique solution *q* on (0, 1) to the equation [37]

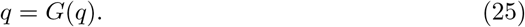

Branching processes which go extinct are often referred to as *mortal* branching processes [5], and in epidemiological contexts the terminology *minor outbreak* is used.

To effectively control an outbreak (in the sense of guaranteeing that all outbreaks go extinct), we need to force the effective reproductive ratio *R*_*e*_ below one. Consider a control measure that reduces the average number of susceptible contacts made during an infectious period to *N*_*c*_ *< N*. This measure reduces the mean of the Poisson contact distribution while leaving the mean of the beta transmission probability intact at its value of *λ/N*. Under this control measure the effective reproductive ratio is

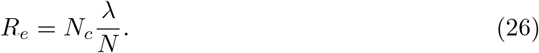

Thus, *R*_*e*_ is below one when *N*_*c*_ *< N/λ*. Such a control measure can be achieved by instituting quarantine measures or school/work closures to reduce the number of contacts made, or through a vaccination program which reduces the number of susceptible contacts made. In the latter case, we observe that at the critical level of vaccination *N* − *N*_*c*_ = *N* (1 − 1*/λ*), giving us the critical vaccination formula commonly found for compartmental models [38].

### 4 Minor outbreak size calculations

The probability that a mortal branching process with *m* initial particles attains a total size of *Z* is given by the joint probability that the *Z* particles generate *Z* − *m* particles between them (i.e. there are as many birth events produced as there are progeny in the entire process), scaled through by a factor of *m/Z*. The outbreak size probability *P* (*Z* = *z*|*m*) in our branching process model is thus given by

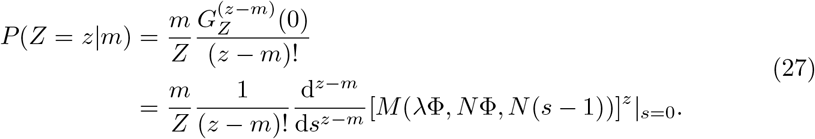

The *i*th derivative of the confluent hypergeometric function *M* (*a, b, z*) is given by [36]:

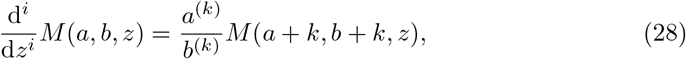

and so

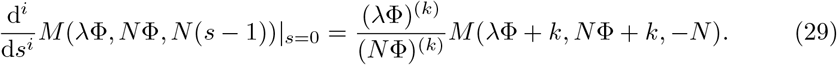

Using the Leibniz rule:

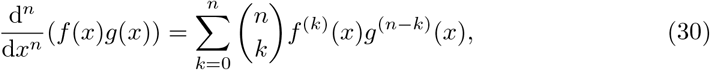

Equation 27 becomes

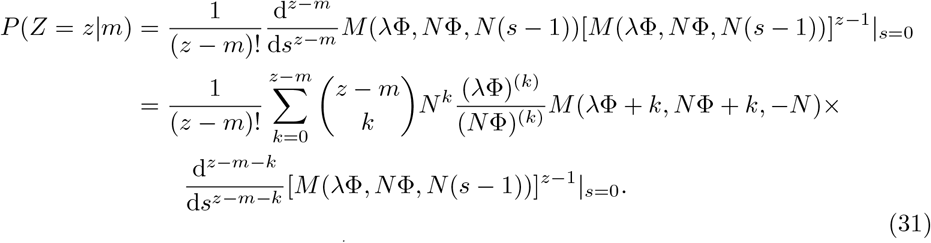

Derivatives of the form 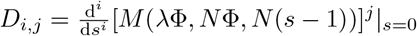 can be calculated recursively using the Liebniz rule:

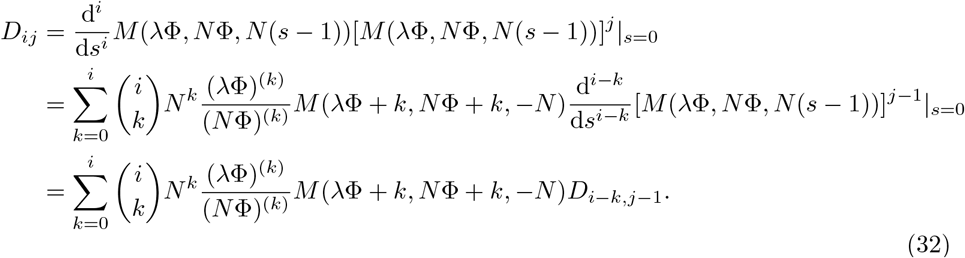

The *j*th column of the matrix ***D*** = (*D*_*i,j*_) is expressed in terms of the *j* − 1th row. To initiate the recursive process, we note that *D*_*i*,1_ is just the *i*th derivative of the confluent hypergeometric function *M* (*λ*Φ, *Nφ, N* (*s* − 1) evaluated at *s* = 0,

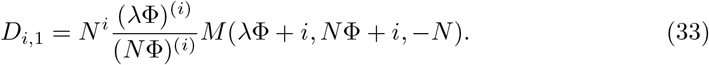

To calculate the probability of obtaining an outbreak of size *z* from *m* index cases, we need to calculate the (*z* − *m*) × *z* submatrix ***D***^*m,z*^ of ***D***. This submatrix contains all the submatrices necessary for calculating all the outbreak size probabilities up to *P* (*Z* = *z*|*m*), and so we effectively obtain the entire distribution up to *z*.

### 5 Major outbreak attack rate

The attack rate of a major epidemic in a large population is approximately Gaussian with mean *Z*_∞_, which is the unique solution to the equation [37, Chapter 4]

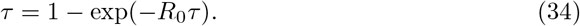

The standard deviation is given by the formula [37, Chapter 4]

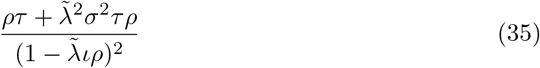

where *τ* = *Z*_∞_, *ρ* = 1 − *τ*. This formula describes a Poisson process of fixed intensity 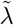 across the population, which takes place over a randomly distributed infectious period with expectation *ι* and variance *σ*^2^. Although our model does not make any assumptions about the distribution of infectious period, it is statistically equivalent to an epidemic with intensity *N* over a beta-distributed infectious period with mean 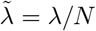 and variance 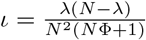. The standard deviation of the major epidemic attack rate is thus

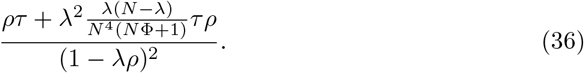

Since the major epidemic attack rate distribution is conditional on not going extinct in the early branching stages of the epidemic, it needs to be scaled by a factor of (1 − *q*) to give the absolute probabilities of attaining these sizes.

### 6 Likelihood calculations using outbreak size data

By fixing an average number of contacts *N*, the beta-Poisson model makes explicit assumptions about underlying contact structure. This means that when fitting the model to outbreak size data we require that all of the outbreaks be from the same underlying population (or a set of populations with similar contact behaviour). For a dataset containing *Z* cases attributable to *m* importations, a maximum likelihood estimator for *λ* is given by [5]

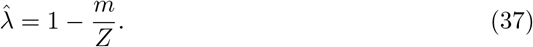

This formula is accurate only when none of the importations have resulted in major outbreaks. Becker [5] outlines the calculation of an MLE when some of the outbreaks are in fact major, but this only works when the underlying offspring distribution is a power series distribution.

Using the minor outbreak size distribution to perform likelihood calculations is very inefficient, since we need to calculate all outbreak size probabilities up to the maximum in our dataset. Intuitively we would expect this distribution to be heavy-tailed, containing large outbreaks with big gaps between the outbreak sizes. This means we need to calculate lots of outbreak size probabilities which we do not directly use. One possible improvement when the underlying population is relatively small is to use the minor outbreak size distribution for outbreak sizes below a certain threshold, and the normally distributed major attack rate distribution for outbreak sizes above that threshold. Because the branching process approximation begins to diverge from the standard epidemic model when the epidemic size reaches the square root of the population size [39], a natural threshold for a population of size *M* is 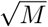. The cumulative outbreak size probability up to 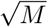 may be substantially smaller than the extinction probability *q*, and so we denote this cumulative probability *q*_*M*_. This is the probability of a minor outbreak in a population of size *M*, using the definition in terms of divergence from the standard epidemic model. The normally distributed major attack rate distribution is scaled by (1 − *q*_*M*_) rather than (1 − *q*). Andersson and Britton [37] suggest that the normal approximation for major outbreaks is reasonably accurate for populations of around size 100, and so this fitting procedure is most useful for populations in the low hundreds, becoming less useful as 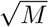 grows and we need to carry out larger and larger matrix calculations.

### 7 Fitted parameter values

Parameter fits for the negative binomial and ZIP models are provided in Tables A and B. Fits and confidence intervals for the parameter *λ* in the Poisson and geometric models are identical to those for *λ* in the negative binomial and beta-Poisson models and so are not listed here.

**Table A.** Maximum likelihood estimates of negative binomial model parameters by dataset, 95% confidence intervals in parentheses.

| Dataset | $\lambda$ | $\theta$ |
| --- | --- | --- |
| Plague | 1.32 (0.88, 1.82) | 0.92 (0.12, 1.98) |
| Mpox | 0.3 (0.22, 0.4) | 0.52 (0.1, 1.05) |
| Ebola, Nigeria 2014 | 0.95 (0.15, 2.3) | 6.89 (0.0, 24.55) |
| Ebola, Guinea 2014 | 0.95 (0.61, 1.36) | 5.26 (2.66, 8.77) |
| SARS, Singapore 2003 | 0.79 (0.36, 1.38) | 8.5 (2.07, 18.87) |
| MERS, South Korea 2015 | 1.05 (0.2, 2.31) | 28.18 (1.91, 77.98) |
| MERS, Saudi Arabia 2015 | 0.96 (0.42, 1.67) | 1.33 (0.0, 4.0) |
| Norovirus, Netherlands 2012 | 0.93 (0.61, 1.3) | 0.56 (0.0, 1.39) |

**Table B.** Maximum likelihood estimates of zero-inflated Poisson model parameters by dataset, 95% confidence intervals in parentheses.

| Dataset | $\tilde{\lambda}$ | $\sigma$ |
| --- | --- | --- |
| Plague | 1.87 (1.15, 2.6) | 0.29 (0.05, 0.48) |
| Mpox | 0.72 (0.37, 1.09) | 0.58 (0.28, 0.72) |
| Ebola, Nigeria 2014 | 3.71 (0.15, 8.33) | 0.74 (0.0, 0.9) |
| Ebola, Guinea 2014 | 3.24 (2.12, 4.46) | 0.71 (0.62, 0.78) |
| SARS, Singapore 2003 | 4.0 (1.63, 6.8) | 0.8 (0.73, 0.86) |
| MERS, South Korea 2015 | 8.7 (1.47, 18.67) | 0.88 (0.82, 0.92) |
| MERS, Saudi Arabia 2015 | 1.71 (0.61, 3.13) | 0.44 (0.0, 0.68) |
| Norovirus, Netherlands 2012 | 1.31 (0.73, 1.93) | 0.28 (0.0, 0.5) |

### 8 Sensitivity analysis

To better understand the fitting behaviour of the beta-Poisson model we calculate one-dimensional cross-sections of the likelihood surface associated with each dataset. In each case we calculate each of the beta-Poisson parameters *λ*, Φ, and *ν* over a range of values with the other two parameters fixed at their MLEs. These cross-sections are plotted in Fig A.

**Fig A.**
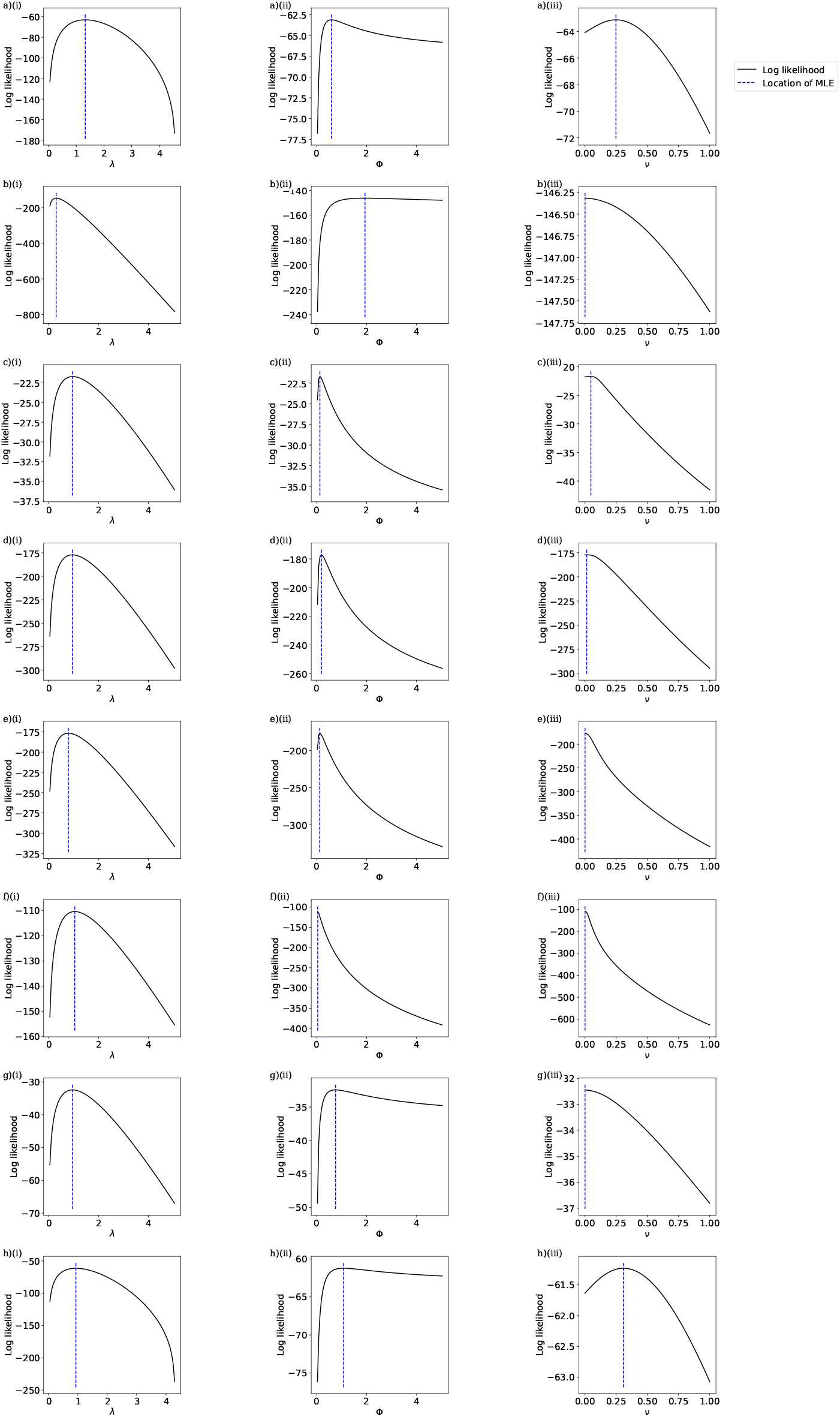
Log-likelihood curves of the beta-Poisson model parameters by dataset. Row a) plague; row b) mpox; row c) Ebola, Nigeria 2014; row d) Ebola, Guinea 2014; row e) SARS, Singapore 2003; row f) MERS, South Korea 2015; row g) MERS, Saudi Arabia 2015; row h) norovirus, the Netherlands 2012; column (i) log-likelihood of *λ* values with Φ and *ν* fixed at MLEs; column (ii) log-likelihood of Φ values with *λ* and *ν* fixed at MLEs; column (iii) log-likelihood of *ν* values with *λ* and Φ fixed at MLEs.

The plots in the middle column of Fig A reveal substantially different fitting behaviour for the parameter Φ depending on the level of overdispersion seen in the dataset in question. Fig A a)(ii), b)(ii), g)(ii), and h)(ii), corresponding respectively to the plague, Mpox, Saudi Arabian MERS and norovirus datasets (all with low levels of overdispersion) all show a comparatively gentle decrease in likelihood as Φ increases away from its MLE, whereas Fig A c)(ii), d)(ii), e)(ii), and f)(ii), corresponding to the more overdispersed Nigerian Ebola, Guinean Ebola, SARS, and South Korean MERS datasets, all display a much more defined peak around the MLE. The third column shows a high likelihood assigned to *ν* = 0 for all the datasets, supporting our finding that the beta-Poisson is unable to offer a substantial improvement in likelihood over the negative binomial model.

